# Using rapid response system trigger clusters to characterize patterns of clinical deterioration among hospitalized adult patients

**DOI:** 10.1101/2023.02.06.23285560

**Authors:** Rebecca J. Piasecki, Elizabeth A. Hunt, Nancy Perrin, Erin M. Spaulding, Bradford Winters, Laura Samuel, Patricia M. Davidson, Nisha Chandra Strobos, Matthew Churpek, Cheryl R. Himmelfarb, the American Heart Association’s Get With The Guidelines^®^-Resuscitation Investigators

**Affiliations:** Johns Hopkins University; University of Wollongong Australia; University of Wisconsin-Madison

## Abstract

**Background:** Many rapid response system (RRS) events are activated using multiple triggers. However, the patterns in which RRS triggers co-occur to activate the medical emergency team (MET) to respond to RRS events is unknown. The purpose of this study was to identify and describe the patterns (RRS trigger clusters) in which RRS triggers co-occur when used to activate the MET and determine the association of these clusters with outcomes using a sample of hospitalized adult patients.

**Methods:** RRS events among adult patients from January 2015 to December 2019 in the Get With The Guidelines-Resuscitation registry’s MET module were examined (n=134,406). A combination of cluster analyses methods was performed to group patients into RRS trigger clusters based on the triggers used to activate their RRS events. Pearson’s chi-squared and ANOVA tests were used to examine differences in patient characteristics across RRS trigger clusters. Multilevel logistic regression was used to examine the associations between RRS trigger clusters and outcomes following RRS events.

**Results:** Six RRS trigger clusters were identified in the study sample. The RRS triggers that predominantly identified each cluster were as follows: tachypnea, new onset difficulty in breathing, and decreased oxygen saturation (Cluster 1); tachypnea, decreased oxygen saturation, and staff concern (Cluster 2); respiratory depression, decreased oxygen saturation, and mental status changes (Cluster 3); tachycardia and staff concern (Cluster 4); mental status changes (Cluster 5); hypotension and staff concern (Cluster 6). Significant differences in patient characteristics were observed across RRS trigger clusters. Patients in Clusters 3 and 6 were associated with an increased likelihood of in-hospital cardiac arrest (IHCA [p<0.01]), while Cluster 4 was associated with a decreased likelihood of IHCA (p<0.01). All clusters were associated with an increased risk of mortality (p<0.01).

**Conclusions:** We discovered six novel RRS trigger clusters with differing relationships to adverse patient outcomes following RRS events. RRS trigger clusters may prove crucial in clarifying the associations between RRS events and adverse outcomes and may aid in clinician decision-making during RRS events.

## Introduction

Since the Institute for Healthcare Improvement in the United States (US) first recommended the use of rapid response systems (RRSs) to improve care for deteriorating inpatients on general hospital wards, they have been widely implemented in many countries, including the US (1-5). RRSs consist of afferent limbs – detecting and activating responses to acutely deteriorating patients – and efferent limbs – providing interventions via specialized teams of clinicians to stabilize deterioration processes and avoid adverse outcomes. These teams that function within RRSs provide urgent interventions for clinically deteriorating patients with the goal of avoiding adverse outcomes, such as in-hospital cardiac arrest (IHCA) and mortality (6-9). While these teams have established standardized nomenclature based on the clinicians involved (e.g., nurse led teams are known as Rapid Response Teams, or RRTs, and Medical Emergency Teams, or METs, also include physicians on the team), for simplicity, here we will refer to all version of these teams as METs. RRSs also include administrative components, such as data management and analysis (7).

RRSs decrease the incidence of non-intensive care unit (non-ICU) IHCAs in adults by up to 33% (8,10-12), but their impacts on other adverse outcomes are not clear (3-5,8,10-13). Increased understanding of key aspects of RRSs, such as RRS triggers, may improve RRS performance and clarify the impact RRSs have on a variety of adverse outcomes for hospitalized patients. RRS triggers are observations or reports of acute changes in a patient’s health that indicate the patient may be experiencing a serious life-threatening clinical deterioration requiring urgent interventions and are used to activate the MET. Up to 44% of adult RRS events are activated using multiple RRS triggers (14,15), and adult patients whose RRS events were activated using two or more triggers have been associated with higher incidences of IHCA and hospital mortality compared to those whose RRS events were activated by single triggers (16,17). Understanding of the patterns in which multiple RRS triggers co-occur may prove crucial in optimizing the care of hospitalized patients with multiple RRS triggers present at the time of RRS activation. However, these patterns have yet to be explored.

The patterns in which RRS triggers occur together resulting in a RRS event activation can be described as RRS trigger clusters. The concept of RRS trigger clusters is derived from symptom clusters – groups of distinct yet related symptoms that tend to occur together in a given disease process – and can be used to better understand disease processes and outcomes and help clinicians more effectively intervene while at the bedside during a RRS event (18). RRS trigger clusters are therefore groups of distinct, yet related individual RRS triggers that tend to co-occur during clinical deterioration processes leading to MET activation. Understanding how RRS trigger clusters are associated with outcomes can provide a more complete picture of clinical deterioration processes and can help researchers and clinicians develop strategies to optimize RRSs and further reduce the incidence of adverse outcomes for hospitalized patients.

### Purpose

The purpose of this study was to identify RRS trigger clusters among a sample of hospitalized adult patients who experienced a RRS event in the US and examine how those clusters were related to patient outcomes following RRS events.

## Methods

### Data Source

This cross-sectional study used the de-identified data stored in the MET module of the American Heart Association’s Get With The Guidelines® – Resuscitation (GWTG-R) registry. Established in 1999, this registry was created as a quality improvement registry by which participating hospitals could benchmark their own performance and compare their resuscitation practices and outcomes to other participating hospitals. The MET module collects data on any patient, visitor, employee, or hospital staff member who experiences a RRS event at a participating site (19-21). Despite its name, the MET module is inclusive of many RRS data elements, not just the response team. The rigorous training and data collection procedures used to gather data for this registry have been described previously (17,20).

An IRB waiver of consent was obtained for this study.

### Study Sample

Records for adult patients who were hospitalized between January 2015 through December 2019 were used. Since we were interested in examining the patterns in which multiple RRS triggers were used to activate METs, only RRS events activated using more than one RRS trigger were considered. A total of 275,062 initial RRS events were identified during the study period. Approximately 49% (n=134,406) of those RRS events had at least two RRS triggers documented as the reason for activation of the MET and were included in the sample.

Adult patients were defined as those aged 18 years or older at the time of their initial RRS event. We excluded any hospital employees, staff members, visitors, outpatients, or patients less than 18 years old at the time of their initial RRS event. Only data from the initial RRS event in each admission (or index event) was analyzed. Data from any additional RRS events that occurred during the same admission were excluded.

### Study Variables

#### RRS Trigger Variables

Table 1 lists the individual RRS triggers collected in the GWTG-R MET module. Using standardized training to extract information on RRS events from patients’ electronic medical records, each RRS trigger had been assessed by GWTG-R data abstractors dichotomously – as either present or not present – for each patient in the study sample.

**Table 1.**
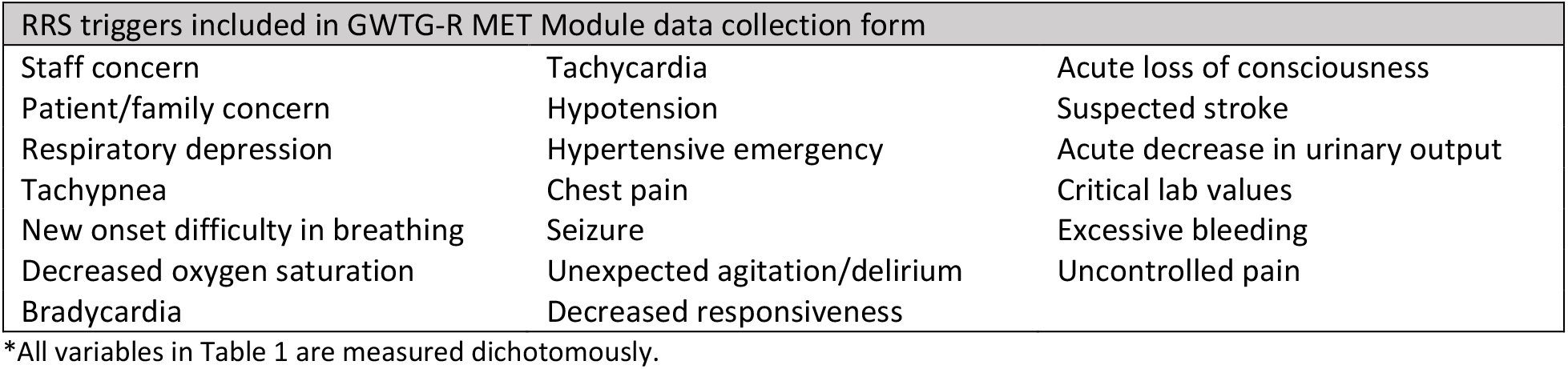
Potential RRS triggers cited as reason for activating the MET*.

| RRS triggers included in GWTG-R MET Module data collection form |  |  |
| --- | --- | --- |
| Staff concern | Tachycardia | Acute loss of consciousness |
| Patient/family concern | Hypotension | Suspected stroke |
| Respiratory depression | Hypertensive emergency | Acute decrease in urinary output |
| Tachypnea | Chest pain | Critical lab values |
| New onset difficulty in breathing | Seizure | Excessive bleeding |
| Decreased oxygen saturation | Unexpected agitation/delirium | Uncontrolled pain |
| Bradycardia | Decreased responsiveness |  |

\*All variables in Table 1 are measured dichotomously.

#### Covariates and Outcome Variables

Several outcomes of interest were identified based on previous RRS research (7-11), with hospital mortality chosen as the primary outcome of interest. Secondary outcomes of interest included: IHCA, acute respiratory compromise (or ARC, which is defined as inadequate respiratory effort necessitating emergent intervention and assisted ventilation), transfer to critical care, limitations placed on code status (defined as “Do Not Resuscitate” or similar orders limiting scope of treatment placed either during or after the RRS event), and any serious adverse event (defined as any combination of the other outcomes of interest occurring during the same admission as the RRS event). Several patient characteristics were considered and controlled for in the analyses for this study including age at time of RRS event, sex, race, ethnicity, primary admitting diagnosis, time of RRS event (event occurred on day versus night shift, or on a weekday versus a weekend), and specialty care received during admission prior to RRS event. Further details describing the operational definitions of the outcome variables and patient characteristic variables are given in Supplemental Table 1.

### Statistical Analysis

#### Cluster Analyses

We used descriptive statistics (frequencies, proportions, percentages) to characterize patients with multiple RRS triggers among each of the 20 RRS triggers in the study sample. Since triggers occurring with a frequency of less than 3% do not have sufficient variability to contribute to the cluster analyses, we excluded any RRS trigger occurring in less than 3% of the sample from subsequent analyses.

A combination of cluster analysis methods was used to group the entire study sample into mutually exclusive RRS trigger clusters. K-means cluster analysis is a powerful and widely used method for determining data clusters; however, it requires *a priori* knowledge of how many clusters are expected to form in a given dataset (29). Hierarchical cluster analysis is a separate method that can be used to help determine the optimal number of clusters in a dataset and can be used in conjunction with k-means cluster analysis (23,29).

The sample was randomly split into a training sample (approximately 67%) and a testing sample (approximately 33%). Hierarchical cluster analysis was performed on the RRS trigger data of training sample and the results were reviewed by the research team to determine the ideal number of clusters to be used in the k-means cluster analyses. Using the ideal number of clusters identified in the hierarchical cluster analysis, k-means cluster analyses were then performed on the training and testing samples to determine patient membership into RRS trigger clusters. Discriminant analyses were performed on both samples with the RRS triggers as predictors of cluster membership to assess how well the identified clusters accounted for the variability in the data (22). Since the inclusion criteria required that at least two RRS triggers be documented as being used to activate a RRS event, no RRS trigger data was documented as missing for any patient included in the cluster analyses. The entire study sample was grouped into the RRS clusters at the completion of the cluster analyses, and subsequent analyses were performed on the entire study sample. The prevalence of RRS triggers in each cluster were described, and any trigger present for at least half of the patients in a cluster was determined to be a characterizing or predominant trigger for that cluster.

All cluster analyses were performed using SPSS version 24 (IBM Corp, Armonk, NY, 2016).

#### Multilevel Logistic Regression Models

To examine how patient characteristics varied across RRS trigger clusters, we first used descriptive statistics (means, standard deviation, frequencies, proportions) to characterize the patient characteristic variables within each RRS trigger cluster. Approximately 5% of the study sample was missing at least one value for at least one patient characteristic variable; therefore, multiple imputation was considered appropriate for the study sample (using ten imputed datasets) and was used to handle missing values for the patient characteristics in the analyses. Pearson’s chi-squared and ANOVA tests were used to examine differences in patient characteristics across RRS trigger clusters for the entire study sample.

Multilevel logistic regression models with patients nested within hospitals, adjusting for patient characteristics (age, sex, race, ethnicity, illness category (based on admission diagnosis), discharged from an intensive care unit (ICU) any time prior to the initial RRS event, discharged from an ICU within 24 hours prior to the initial RRS event, discharged from the emergency department within 24 hours prior to the initial RRS event, received sedation within 24 hours prior to the initial RRS event, and timing of the initial RRS event), were then used to determine the associations between RRS trigger clusters and patient outcomes. Frequencies and proportions were used to characterize outcomes (hospital mortality, IHCA, ARC, transfer to critical care, limitations placed on code status, and any serious adverse event) for each RRS trigger cluster. Separate multilevel logistic regression models with patients nested within hospitals were conducted for the outcomes of interest related to RRS events. Patient characteristics were included as covariates in the first level of the models and hospital clustering effects were accounted for in the second level.

The above statistical tests were performed using Stata version 16.1 (StataCorp, College Station, TX, 2019).

## Results

### Study Sample Characteristics

The mean age in the study sample was 66 years (SD=17), and 51% of the sample was female. Patients who identified as White comprised 73.0% of the sample, while 23.8% of the sample identified themselves through another racial identity (labeled as “All other races” in this study). Approximately 4.6% of the study sample identified as Hispanic compared to 95.4% as non-Hispanic.

### Cluster Analyses

Overall, 13 of the 20 RRS triggers assessed were present in at least 3% of the RRS events in the data set (see Table 2 for details on the prevalence of these RRS triggers). The seven RRS triggers excluded from the cluster analyses due to a prevalence of less than 3% in the study sample were: patient or family concern, chest pain, acute decrease urinary output, critical lab abnormality, risk factor score, excessive bleeding, and uncontrolled pain. Using the results of the hierarchical cluster analysis, the research team examined 4, 5, 6, and 7 cluster solutions for clinical interpretability and significance of each possible solution. A consensus was reached that the 6-cluster solution would be optimal to inform the subsequent k-means clusters analyses on the training (n=88,431) and testing (n=45,975) samples. Discriminant analyses performed to assess for goodness-of-fit of the final RRS trigger clusters resulted in 93.4% of patients into the same clusters as the cluster analyses in the training sample (with a canonical correlation coefficient of 0.946), and 94.3% of patients into the same clusters as the cluster analyses in the testing sample (with a canonical correlation coefficient of 0.949). This indicates a high degree of agreement in the classification of patients to clusters between the cluster analyses and the discriminant analyses and validates the results of the cluster analyses.

**Table 2.** Prevalence of RRS triggers used in cluster analysis in the entire study sample.

| RRS Trigger | Study Sample<br>(n=134,406)<br>No. (%) |
| --- | --- |
| Mental status change | 55,697 (41.4) |
| Staff concern | 55,247 (41.1) |
| Decreased oxygen saturation | 41,412 (30.8) |
| New onset difficulty in breathing | 33,160 (24.7) |
| Tachycardia | 33,020 (24.6) |
| Tachypnea | 28,967 (21.6) |
| Hypotension | 28,206 (21.0) |
| Acute loss of consciousness | 8,577 (6.4) |
| Respiratory depression | 8,082 (6.0) |
| Bradycardia | 6,706 (5.0) |
| Suspected acute stroke | 6,588 (4.9) |
| Hypertensive emergency | 6,115 (4.5) |
| Seizure | 5,934 (4.4) |

Clusters 1, 2, and 3 were all primarily defined by at least one respiratory-associated RRS trigger (Table 3). Cluster 1 was exclusively characterized by RRS triggers related to respiratory deterioration – tachypnea, new onset difficulty in breathing, and decreased oxygen saturation. Cluster 2 was primarily characterized by tachypnea, decreased oxygen saturation, and staff concern. Of note, no RRS triggers related to cardiac, circulatory, and neurological issues were common in either Cluster 1 or 2, and new onset difficulty in breathing was not common in Cluster 2. Cluster 3 was predominantly defined by respiratory depression, decreased oxygen saturation, and mental status changes. No cardiac or circulatory RRS triggers were common in Cluster 3. Cluster 4 was predominantly characterized by tachycardia and staff concern, with low frequencies of respiratory or neurological RRS triggers. Cluster 5 was predominantly characterized only by mental status changes. To clarify, this does not mean that the patients grouped into Cluster 5 only had mental status changes as their sole RRS trigger, but rather, this was the only RRS trigger that a majority of the patients in this cluster had in common. Finally, Cluster 6 was primarily characterized by hypotension and staff concern, with no common respiratory or neurological RRS triggers identified. Additional details of the prevalence of RRS triggers in each cluster in the study sample are given in Table 3.

**Table 3.** Percentages of RRS triggers present across RRS trigger clusters for the entire study sample.

| RRS triggers (%) | Cluster<br>(No. of patients) |  |  |  |  |  |
| --- | --- | --- | --- | --- | --- | --- |
|  | 1<br>(n=28,205) | 2<br>(n=15,750) | 3<br>(n=7,298) | 4<br>(n=18,786) | 5<br>(n=41,372) | 6<br>(n=22,995) |
| Respiratory depression | 1 | 1 | 100 | 1 | 0 | 1 |
| Tachypnea | 49 | 50 | 6 | 26 | 2 | 4 |
| New onset difficulty in breathing | 100 | 0 | 17 | 6 | 4 | 4 |
| Decreased oxygen saturation | 60 | 84 | 45 | 3 | 11 | 12 |
| Bradycardia | 1 | 3 | 8 | 1 | 3 | 17 |
| Tachycardia | 25 | 31 | 10 | 100 | 4 | 0 |
| Hypotension | 4 | 3 | 14 | 36 | 0 | 82 |
| Hypertensive emergency | 6 | 12 | 2 | 7 | 3 | 0 |
| Mental status change | 10 | 8 | 52 | 13 | 91 | 33 |
| Acute loss of consciousness | 1 | 3 | 12 | 2 | 9 | 12 |
| Seizure | 1 | 2 | 2 | 2 | 11 | 1 |
| Suspected acute stroke | 0 | 1 | 1 | 1 | 14 | 1 |
| Staff concern | 32 | 53 | 33 | 51 | 33 | 53 |

### Patient Characteristic Differences between Clusters

We found statistically significant differences between the RRS trigger clusters across all patient characteristics examined (Table 4). For Cluster 4, the mean age (63 years) was lower than all other RRS trigger clusters (range 66 to 68 years), and patients identifying under “All other races” accounted for more than a quarter of the patients in Clusters 4, 5, and 6 as compared to Clusters 1, 2 and 3 (22% or less). Larger proportions of the patients in Clusters 4 and 6 had cardiac admitting diagnoses (29% and 29%, respectively) versus non-cardiac admitting diagnoses when compared to the other RRS trigger clusters (23% or less). Compared to the other RRS trigger clusters, fewer patients in Clusters 5 and 6 were discharged from an ICU prior to their initial RRS event, while more patients in Clusters 3 and 6 received sedation within the 24 hours prior to their initial RRS event. The initial RRS event for patients in Cluster 5 (56%) more often occurred during the day shift as compared to the other RRS trigger clusters (54% or less).

**Table 4.** Patient characteristics across RRS trigger clusters for the entire study sample.

| Cluster<br>(No. of patients) | 1<br>(N=28,205) | 2<br>(N=15,750) | 3<br>(N=7,298) | 4<br>(N=18,786) | 5<br>(N=41,372) | 6<br>(N=22,995) | p-<br>value |
| --- | --- | --- | --- | --- | --- | --- | --- |
| No. of Patients<br>(% of Total) | *n*<br>(%) | *n*<br>(%) | *n*<br>(%) | *n*<br>(%) | *n*<br>(%) | *n*<br>(%) |  |
| Age (mean ± SD) | 67 ± 16 | 67 ± 17 | 68 ± 16 | 63 ± 18 | 66 ± 17 | 67 ± 16 | <0.001 |
| Race |  |  |  |  |  |  | <0.001 |
| White | 21,384<br>(78.0) | 12,000<br>(79.0) | 5,604<br>(80.2) | 13,327<br>(73.3) | 29,313<br>(73.1) | 16,502<br>(74.2) |  |
| All other races | 6,047<br>(22.0) | 3,189<br>(21.0) | 1,387<br>(19.8) | 4,848<br>(26.7) | 10,808<br>(26.9) | 5,752<br>(25.9) |  |
| Ethnicity |  |  |  |  |  |  | <0.001 |
| Hispanic | 1,220<br>(4.3) | 668<br>(4.2) | 322<br>(4.4) | 971<br>(5.2) | 1,928<br>(4.7) | 1,095<br>(4.8) |  |
| Non-Hispanic | 26,985<br>(95.7) | 15,082<br>(95.8) | 6,976<br>(95.6) | 17,815<br>(94.8) | 39,444<br>(95.3) | 21,900<br>(95.2) |  |
| Sex |  |  |  |  |  |  | <0.001 |
| Male | 14,142<br>(50.1) | 8,116<br>(51.5) | 3,536<br>(48.5) | 9,410<br>(50.1) | 19,440<br>(47.0) | 11,157<br>(48.5) |  |
| Female | 14,062<br>(49.9) | 7,633<br>(48.5) | 3,762<br>(51.6) | 9,375<br>(49.9) | 21,929<br>(53.0) | 11,833<br>(51.5) |  |
| Illness Category |  |  |  |  |  |  | <0.001 |
| Medical | 24,446<br>(86.7) | 13,313<br>(84.6) | 6,002<br>(82.3) | 15,309<br>(81.5) | 34,673<br>(83.9) | 18,553<br>(80.7) |  |
| Surgical | 3,754<br>(13.3) | 2,429<br>(15.4) | 1,294<br>(17.7) | 3,475<br>(18.0) | 6,667<br>(16.1) | 4,435<br>(19.3) |  |
| Cardiac | 6,175<br>(21.9) | 3,183<br>(20.2) | 1,712<br>(23.5) | 5,358<br>(28.5) | 7,161<br>(17.3) | 6,779<br>(29.5) | <0.001 |
| Non-Cardiac | 22,025<br>(78.1) | 12,559<br>(79.8) | 5,584<br>(76.5) | 13,426<br>(71.5) | 34,179<br>(82.7) | 16,209<br>(70.5) |  |
| Discharged from ICU any time prior to RRS event |  |  |  |  |  |  |  |
| Yes | 4,868<br>(17.3) | 2,672<br>(17.0) | 1,112<br>(15.2) | 2,629<br>(14.0) | 5,464<br>(13.2) | 2,952<br>(12.8) | <0.001 |
| No | 23,336<br>(82.7) | 13,076<br>(83.0) | 6,183<br>(84.8) | 16,156<br>(86.0) | 35,880<br>(86.8) | 20,040<br>(87.2) |  |
| Discharged from ICU 24h prior to RRS event |  |  |  |  |  |  |  |
| Yes | 1,521<br>(5.4) | 823<br>(5.2) | 325<br>(4.5) | 698<br>(3.7) | 1,476<br>(3.6) | 699<br>(3.0) | <0.001 |
| No | 26,665<br>(94.6) | 14,921<br>(94.8) | 6,969<br>(95.5) | 18,080<br>(96.3) | 39,861<br>(96.4) | 22,289<br>(97.0) |  |
| Discharge from ED 24h prior to RRS event |  |  |  |  |  |  |  |
| Yes | 6,613<br>(23.5) | 3,561<br>(22.6) | 1,579<br>(21.6) | 4,548<br>(24.2) | 9,600<br>(23.2) | 5,843<br>(25.4) | <0.001 |
| No | 21,590<br>(76.6) | 12,185<br>(77.4) | 5,716<br>(78.4) | 14,236<br>(75.8) | 31,744<br>(76.8) | 17,149<br>(74.6) |  |
| Received sedation 24h prior to RRS event |  |  |  |  |  |  |  |
| Yes | 1,655<br>(5.9) | 943<br>(6.0) | 821<br>(11.3) | 1,578<br>(8.4) | 3,079<br>(7.5) | 2,922<br>(12.7) | <0.001 |
| No | 26,547<br>(94.1) | 14,803<br>(94.0) | 6,472<br>(88.7) | 17,207<br>(91.6) | 38,264<br>(92.6) | 20,069<br>(87.3) |  |
| Timing of RRS Event |  |  |  |  |  |  |  |
| Day (07:00-18:59) | 14,305<br>(50.7) | 7,817<br>(49.6) | 3,813<br>(52.3) | 9,572<br>(51.0) | 23,284<br>(56.3) | 12,509<br>(54.4) | <0.001 |
| Night (19:00-06:59) | 13,900<br>(49.3) | 7,933<br>(50.4) | 3,485<br>(47.8) | 9,214<br>(49.1) | 18,088<br>(43.7) | 10,486<br>(45.6) |  |
| Weekend (19:00 F – 06:59 M) | 9,908 (35.1) | 5,574<br>(35.4) | 2,325<br>(31.9) | 6,528<br>(34.8) | 13,609<br>(32.9) | 7,457<br>(32.4) | <0.001 |
| Weekday (07:00 M – 18:59 F) | 18,297<br>(64.9) | 10,176<br>(64.6) | 4,973<br>(68.1) | 12,258<br>(65.3) | 27,763<br>(67.1) | 15,538<br>(67.6) |  |

### Outcome Differences between Clusters

We also found statistically significant differences between RRS trigger clusters across all outcomes examined (Table 5). For example, patients in Cluster 3 had the highest reported mortality (28%) compared to the other RRS trigger clusters (20% or less). Patients in Cluster 3 also had the highest reported incidences of IHCA (5%) compared to other clusters (2% or less). Additional details regarding differences in adverse outcomes across RRS trigger clusters can be found in Table 5.

**Table 5.**
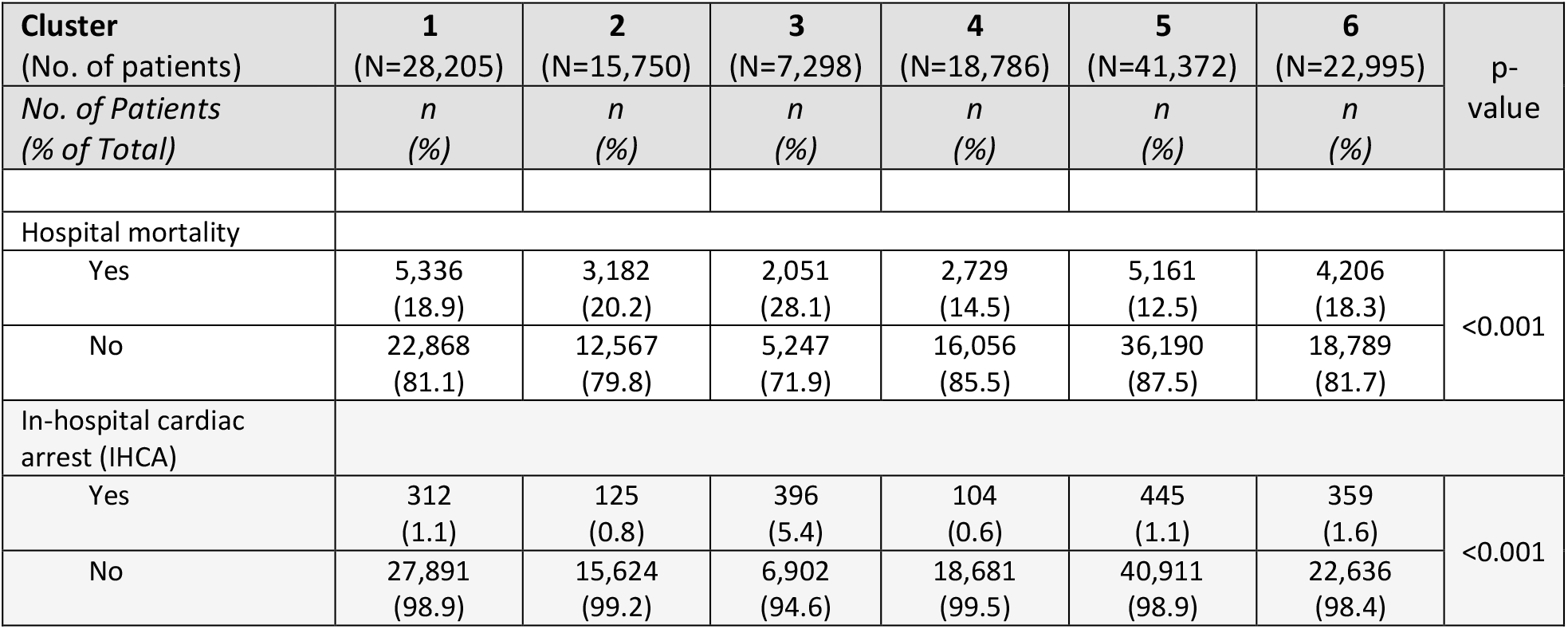
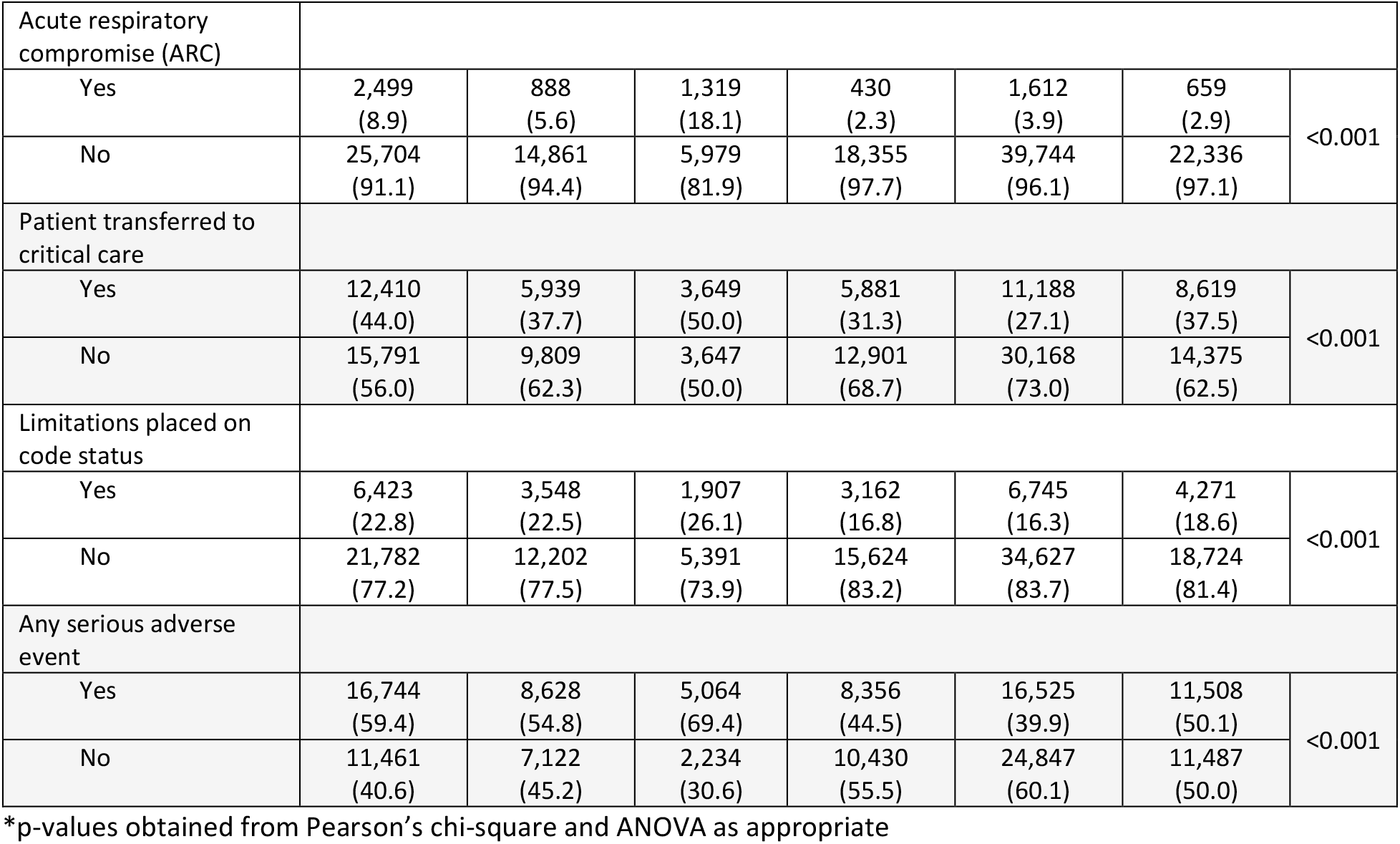
Adverse patient outcomes across RRS trigger clusters for the entire study sample*.

| Cluster<br>(No. of patients) | 1<br>(N=28,205) | 2<br>(N=15,750) | 3<br>(N=7,298) | 4<br>(N=18,786) | 5<br>(N=41,372) | 6<br>(N=22,995) | p-<br>value |
| --- | --- | --- | --- | --- | --- | --- | --- |
| No. of Patients<br>(% of Total) | *n*<br>(%) | *n*<br>(%) | *n*<br>(%) | *n*<br>(%) | *n*<br>(%) | *n*<br>(%) |  |
| Hospital mortality |  |  |  |  |  |  |  |
| Yes | 5,336<br>(18.9) | 3,182<br>(20.2) | 2,051<br>(28.1) | 2,729<br>(14.5) | 5,161<br>(12.5) | 4,206<br>(18.3) | <0.001 |
| No | 22,868<br>(81.1) | 12,567<br>(79.8) | 5,247<br>(71.9) | 16,056<br>(85.5) | 36,190<br>(87.5) | 18,789<br>(81.7) |  |
| In-hospital cardiac<br>arrest (IHCA) |  |  |  |  |  |  |  |
| Yes | 312<br>(1.1) | 125<br>(0.8) | 396<br>(5.4) | 104<br>(0.6) | 445<br>(1.1) | 359<br>(1.6) | <0.001 |
| No | 27,891<br>(98.9) | 15,624<br>(99.2) | 6,902<br>(94.6) | 18,681<br>(99.5) | 40,911<br>(98.9) | 22,636<br>(98.4) |  |
| Acute respiratory compromise (ARC) |  |  |  |  |  |  |  |
| Yes | 2,499<br>(8.9) | 888<br>(5.6) | 1,319<br>(18.1) | 430<br>(2.3) | 1,612<br>(3.9) | 659<br>(2.9) | <0.001 |
| No | 25,704<br>(91.1) | 14,861<br>(94.4) | 5,979<br>(81.9) | 18,355<br>(97.7) | 39,744<br>(96.1) | 22,336<br>(97.1) |  |
| Patient transferred to critical care |  |  |  |  |  |  |  |
| Yes | 12,410<br>(44.0) | 5,939<br>(37.7) | 3,649<br>(50.0) | 5,881<br>(31.3) | 11,188<br>(27.1) | 8,619<br>(37.5) | <0.001 |
| No | 15,791<br>(56.0) | 9,809<br>(62.3) | 3,647<br>(50.0) | 12,901<br>(68.7) | 30,168<br>(73.0) | 14,375<br>(62.5) |  |
| Limitations placed on code status |  |  |  |  |  |  |  |
| Yes | 6,423<br>(22.8) | 3,548<br>(22.5) | 1,907<br>(26.1) | 3,162<br>(16.8) | 6,745<br>(16.3) | 4,271<br>(18.6) | <0.001 |
| No | 21,782<br>(77.2) | 12,202<br>(77.5) | 5,391<br>(73.9) | 15,624<br>(83.2) | 34,627<br>(83.7) | 18,724<br>(81.4) |  |
| Any serious adverse event |  |  |  |  |  |  |  |
| Yes | 16,744<br>(59.4) | 8,628<br>(54.8) | 5,064<br>(69.4) | 8,356<br>(44.5) | 16,525<br>(39.9) | 11,508<br>(50.1) | <0.001 |
| No | 11,461<br>(40.6) | 7,122<br>(45.2) | 2,234<br>(30.6) | 10,430<br>(55.5) | 24,847<br>(60.1) | 11,487<br>(50.0) |  |

\*p-values obtained from Pearson's chi-square and ANOVA as appropriate

### Multilevel Logistic Regression Models

In the multilevel logistic regression models (Table 6), Cluster 5, which was predominantly defined by mental status changes, was used as the reference cluster since it contained the most patients and the lowest mortality of all the clusters (12%). Patients in Clusters 1, 2, and 3 were more likely to experience ARC compared to the reference cluster (Cluster 1: OR=2.70, 95% CI=2.52 to 2.90; Cluster 2: OR=1.56, 95% CI=1.42 to 1.70; and Cluster 3: OR=5.69, 95% CI=5.22 to 6.20), whereas patients in Clusters 4 and 6 were less likely to experience ARC (OR=0.54, 95% CI: 0.48 to 0.60; and OR=0.73, 95% CI: 0.66 to 0.80, respectively). Patients in Clusters 3 and 6 were more likely to experience IHCA compared to the reference cluster (OR=5.20, 95% CI: 4.50 to 6.01; and OR=1.49, 95% CI: 1.29 to 1.72, respectively), but patients in Cluster 4 were less likely to experience IHCA during their admission (OR=0.57, 95% CI: 0.45 to 0.70). Patients in Clusters 1, 2, 3, 4, and 6 were all more likely to have limitations placed on their code status and be transferred to critical care following the RRS event compared to the reference cluster. Table 6 provides additional details of the multilevel logistic regression models for each outcome.

**Table 6.** Associations between RRS trigger clusters and outcomes^+^.

| Cluster | 1 | 2 | 3 | 4 | 5 | 6 |
| --- | --- | --- | --- | --- | --- | --- |
| *Odds Ratio*<br>*(95% Confidence Interval)* | *OR*<br>*(CI)* | *OR*<br>*(CI)* | *OR*<br>*(CI)* | *OR*<br>*(CI)* | *OR*<br>*(CI)* | *OR*<br>*(CI)* |
| Patient Outcomes |  |  |  |  |  |  |
| Hospital mortality | 1.62 **<br>(1.55 - 1.69) | 1.75 **<br>(1.66 - 1.84) | 2.77 **<br>(2.60 - 2.94) | 1.35 **<br>(1.28 - 1.42) | Ref. | 1.70 **<br>(1.62 - 1.78) |
| In-hospital cardiac arrest (IHCA) | 1.04<br>(0.89 - 1.20) | 0.82<br>(0.67 - 1.00) | 5.20 **<br>(4.50 - 6.01) | 0.57 **<br>(0.45 - 0.70) | Ref. | 1.49 **<br>(1.29 - 1.72) |
| Acute respiratory compromise (ARC) | 2.70 **<br>(2.52 - 2.90) | 1.56 **<br>(1.42 - 1.70) | 5.69 **<br>(5.22 - 6.20) | 0.54 **<br>(0.48 - 0.60) | Ref. | 0.73 **<br>(0.66 - 0.80) |
| Patient transferred to critical care | 2.23 **<br>(2.15 - 2.30) | 1.72 **<br>(1.65 - 1.80) | 2.69 **<br>(2.55 - 2.84) | 1.43 **<br>(1.37 - 1.49) | Ref. | 1.73 **<br>(1.67 - 1.80) |
| Limitations placed on code status | 1.44 **<br>(1.38 - 1.50) | 1.47 **<br>(1.41 - 1.55) | 1.83 **<br>(1.72 - 1.94) | 1.24 **<br>(1.18 - 1.30) | Ref. | 1.23 **<br>(1.18 - 1.29) |
| Any serious adverse event | 2.24 **<br>(2.16 - 2.31) | 1.88 **<br>(1.81 - 1.96) | 3.39 **<br>(3.20 - 3.59) | 1.43 **<br>(1.37 - 1.48) | Ref. | 1.62 **<br>(1.56 - 1.67) |

\*\* statistically significant (p<0.01)
<sup>+</sup> Covariates included in multilevel logistical regression models: age, race, ethnicity, sex, illness category, discharge from ICU any time prior to the initial RRS event, discharge from an ICU within 24 hours prior to the initial RRS event, discharge from the emergency department within 24 hours prior to the initial RRS event, received sedation within 24 hours prior to the initial RRS event, timing of the initial RRS event

## Discussion

We found that in the GWTG-R MET module database that approximately 49% (134,406 of 275,062) of index RRS events during the study period had two or more RRS triggers documented as being used to activate a RRS event. Through cluster analyses, we identified an ideal solution of 6 RRS trigger clusters. These trigger clusters were associated with patient characteristics such as admitting diagnosis, recent intensive care unit discharge, use of recent sedation, race, and ethnicity. All clusters were associated with higher likelihood of hospital mortality, transfer to critical care, limitations placed on code status, and any serious adverse event compared with the reference cluster, which was primarily defined by mental status changes. After accounting for patient characteristics and the clustering effects of patients who experienced RRS events within the same hospitals, patients with certain clusters were statistically more likely to experience ARC or IHCA. Of note, the likelihood of any adverse patient outcomes was at least two times higher for the patients in Cluster 3 (primarily characterized by respiratory depression, decreased oxygen saturation, mental status changes) compared to patients with other clusters. Each cluster had its own unique associative profile across the outcomes examined.

Currently, METs are activated using both clinical judgement criteria, such as staff concern, and objective criteria, such as abnormal variations in vital signs (27). These parameters may be aggregated into early warning scores (EWS) or used as single threshold parameters for the purposes of activation (8,12). Previous studies have shown that abnormal variations in vital signs are often present many hours prior to RRS events and adverse patient outcomes, such as IHCA (28). Additionally, recent work suggests that combinations of subjective clinical judgement criteria and multiple objective criteria may improve early detection of acutely deteriorating patients, thus improving RRS interventions and outcomes for these vulnerable patients (17, 27, 28). This study adds to the current understanding of RRSs, especially the afferent limb, by examining the patterns in which subjective and objective RRS triggers co-occur through the conceptualization of RRS trigger clusters.

Given that patients whose RRS events were activated using multiple RRS triggers have been shown to be more likely to experience IHCA or mortality (16,17), it is not surprising that all RRS trigger clusters we identified were associated with high likelihood of hospital mortality (OR=1.35 or greater). The odds ratio for mortality was nearly double among those patients in Cluster 3 (respiratory depression, decreased oxygen saturation, and mental status changes) as compared to the other RRS trigger clusters suggesting it strongly defines severe clinical deterioration. In comparing the defining triggers among each of the clusters, Cluster 3 is unique in that it is the only RRS trigger cluster with prevalent triggers involving more than one organ system – respiratory (100% of patients in the cluster had the respiratory depression trigger present and 45% had the decreased oxygen saturation trigger present) and neurological (52% of patients had the mental status change trigger present). This suggests that when multiple RRS triggers representing multiple organ systems are present, patients may be further along in their deterioration process and/or experiencing a more catastrophic event resulting in a higher risk for hospital mortality, IHCA, or ARC. This finding may be able to be used to guide management at the bedside and inform triage decisions with a goal of targeting a reduction in these adverse outcomes for these patients. This finding should also be taken into consideration in in possible reworking how EWSs function. EWSs often assign “scores” based on how far outside of “normal” parameters each vital sign is. Our findings suggest that these simple methods of aggregating scores are often not sufficient, and that the manner in which these signs and symptoms occur together should also be taken into account when creating EWSs.

Previously reported findings have suggested that staff concern as a RRS trigger is associated with decreased incidence of adverse outcomes following RRS events (24). Here, the defining triggers of Clusters 4 and 6 each included staff concern and one other cardiovascular RRS trigger—tachycardia in Cluster 4 and hypotension in Cluster 6. Cluster 2 also has staff concern as a defining RRS trigger; however, Cluster 2 also had two additional respiratory defining RRS triggers as opposed to Clusters 4 and 6 which each only had one other physiological defining RRS trigger in addition to staff concern. Some interesting differences were noted between these clusters, including how they were associated with IHCA—the patients in cluster 2 had a non-significant lower likelihood of experiencing IHCA, the patients in Cluster 4 were significantly less likely to experience IHCA, and the patients in Cluster 6 were significantly more likely to experience IHCA. While these results contradict some of the previous findings related to the seemingly protective nature of the staff concern trigger, it is important to note that others have found that the use of respiratory triggers to activate RRS events is associated with increased likelihood of adverse outcomes (16,17). This could indicate that while staff concern as a trigger may indicate an opportunity for more aggressive early interventions to avoid adverse outcomes, the physiological triggers that occur with that concern may temper its potentially protective effects. Further study to elucidate the underlying mechanisms behind these differences is paramount.

This study highlights several key areas for future RRS research. Many previous studies have added critical knowledge about the importance of triggers in the afferent RRS limb and how they contribute to recognition of clinical deterioration and outcomes. For example, Shappell and colleagues (2018) examined associations between RRS triggers and outcomes and found that the presence of respiratory triggers were associated with increased risk of mortality (17). While such studies have been important in furthering our understanding of how triggers relate to adverse outcomes, they have tended to examine triggers grouped together by organ system (respiratory triggers, cardiovascular triggers, etc.). Our findings indicate that having a better understanding of RRS trigger clusters that cross organ system boundaries may provide key insights into early recognition of clinical deterioration. We found that almost half of METs are activated using more than one trigger, further highlighting the importance of examining MET activation using multiple triggers and RRS trigger clusters in further detail. Additionally, the RRS trigger clusters identified and defined in this study should be considered for use in bedside decision-making during RRS events especially for the purposes of triage to a higher level of care. Furthermore, RRS trigger clusters may also improve the predictive abilities of early warning scoring systems and machine learning algorithms designed to help clinicians better detect clinical deterioration and improve patient outcomes.

### Limitations

This study has several limitations. First, hospitals participate in the GWTG-R modules voluntarily and they may not be representative of hospitals across the United States. Further study of the use of multiple RRS triggers to activate RRS events, outside of this registry, is warranted. An additional limitation is the widespread availability of clustering algorithms and corresponding lack of a gold standard in their applications in healthcare research. Use of different clustering algorithms may result in different RRS trigger clusters than those described here (30). Another potential limitation is that information regarding patients’ past medical histories and comorbidities are not captured in the GWTG-R MET module. However, patient comorbidities may or may not accurately predict acute clinical deterioration and patient outcomes following serious adverse events (25), so the lack of this patient information may or may not have affected our findings. Additionally, the GWTG-R modules do not report data related to MET composition. How team composition of the efferent limb affects outcomes remains a key area in need of further study (8). Many studies provide limited data on team composition. One recent study found that dedicated interdisciplinary teams are associated with fewer adverse outcomes related to cardiopulmonary resuscitation (26) but similar comparisons are lacking in the RRS literature.

Given that RRS triggers are part of the afferent limb and METs are part of the efferent limb, the lack of information regarding team composition may not have been essential to conceptualizing the trigger clusters. However, future studies examining the spectrum of RRSs should study afferent and efferent limbs together to better understand their relationships with patient outcomes. Just as predictive modeling and machine learning algorithms can provide improved, patient-specific predictions of outcomes, future RRS research should also examine how organization-specific factors are associated with outcomes related to the optimization of RRSs. In examining organization factors along with patient-specific variables across the afferent and efferent limbs of RRSs, we will be able to more fully answer how RRSs can be best implemented.

## Conclusions

This study of a large registry of adult patients experiencing RRS events demonstrated that activation triggers, when occurring in multiples, associate into novel patterns resulting in RRS trigger clusters. All clusters were associated with increased risk of hospital mortality. Cluster 4 was associated with a lower risk of IHCA, while Clusters 3 and 6 were associated with an increased risk of IHCA. RRS trigger clusters could be crucial in guiding bedside care and triage, improving EWSs and prediction algorithms, and helping clinicians and researchers better detect and treat clinical deterioration.

## Data Availability

All data used for this study were obtained from the MET Module of the Get With The Guidelines - Resuscitation registry, owned and administered by the American Heart Association, to whom any requests for data access should be addressed.

## Acknowledgements

The Get With The Guidelines^®^ programs are provided by the American Heart Association. Hospitals participating in the registry submit clinical information regarding the medical history, hospital care, and outcomes of consecutive patients hospitalized for cardiac arrest using an online, interactive case report form and Patient Management Tool™ (IQVIA, Parsippany, New Jersey). IQVIA (Parsippany, New Jersey) serves as the data collection (through their Patient Management Tool – PMT™) and coordination center for the American Heart Association/American Stroke Association Get With The Guidelines® programs. The University of Pennsylvania serves as the data analytic center and has an agreement to prepare the data for research purposes. All participating institutions were required to comply with local regulatory and privacy guidelines and, if required, to secure institutional review board approval. Because data were used primarily at the local site for quality improvement, sites were granted a waiver of informed consent under the common rule. We would like to acknowledge the efforts and contributions of the American Heart Association’s Get With The Guidelines®-Resuscitation Adult Research Task Force members: Anne Grossestreuer PhD; Ari Moskowitz MD; Dana Edelson MD MS; Joseph Ornato MD; Mary Ann Peberdy MD; Matthew Churpek MD MPH PhD; Monique Anderson Starks MD MHS; Paul Chan MD MSc; Saket Girotra MBBS SM; Sarah Perman MD MSCE; and, Zachary Goldberger MD MS.

## Supplemental Material

**Supplemental Table 1.**
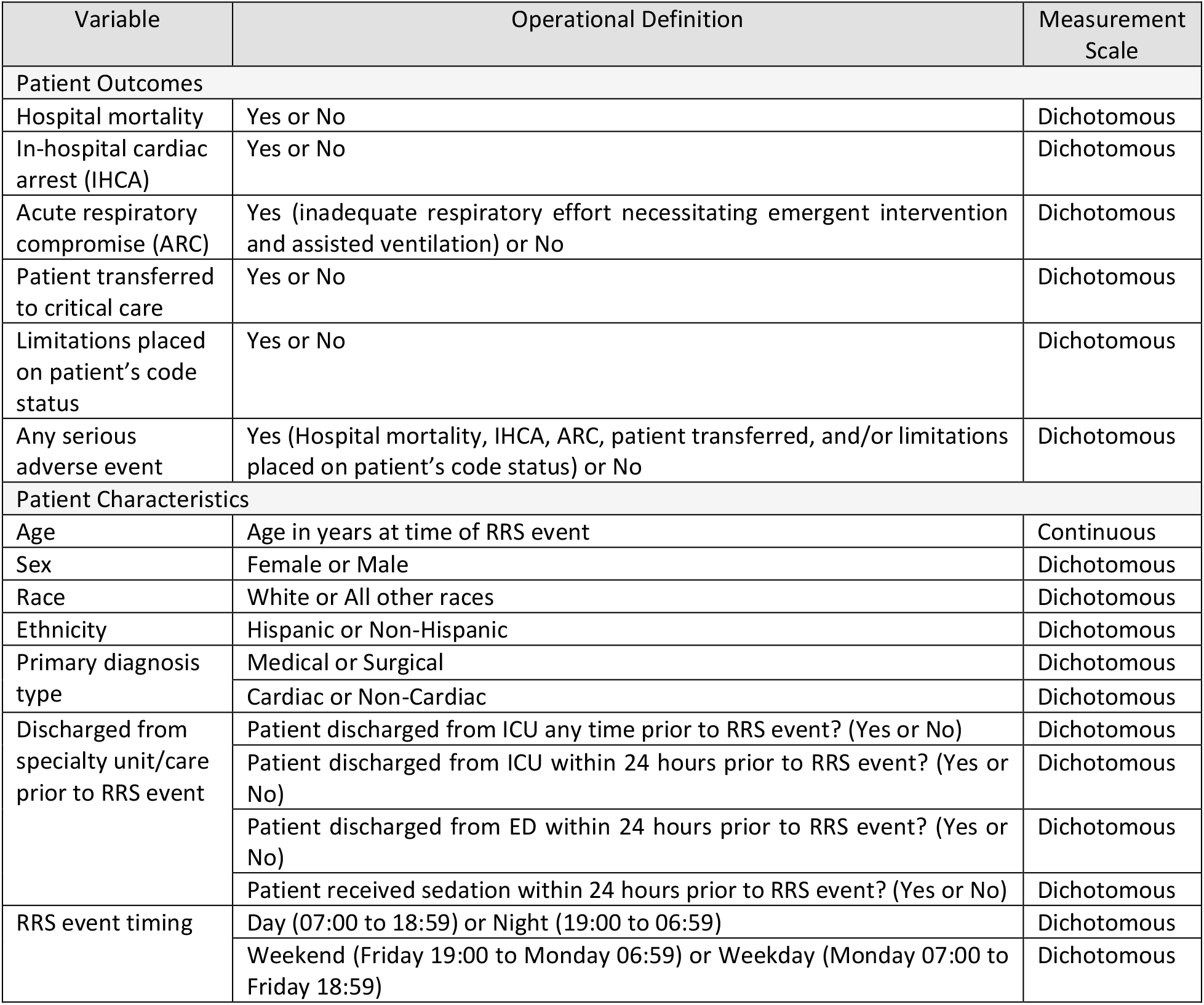
Outcomes and patient characteristics variable definitions.

